# Integrating Mendelian randomization and literature-mined evidence for breast cancer risk factors

**DOI:** 10.1101/2022.07.19.22277795

**Authors:** Marina Vabistsevits, Tim Robinson, Ben Elsworth, Yi Liu, Tom R Gaunt

**Affiliations:** Medical Research Council Integrative Epidemiology Unit at the University of Bristol, University of Bristol, Oakfield 7 House, Oakfield Grove, Bristol, BS8 2BN, UK; Population Health Sciences, Bristol Medical School, University of Bristol, Oakfield 7 House, Oakfield Grove, Bristol, BS8 2BN, UK; Our Future Health, 2 New Bailey, 6 Stanley Street, Manchester, M3 5GS, UK; University of Exeter Medical School, University of Exeter St Luke’s Campus, 79 Heavitree Rd, Exeter, EX2 4TH, UK

**Keywords:** mendelian randomization, genome-wide association study, causal inference, breast cancer, literature-based discovery, knowledge graph, literature mining

## Abstract

Graphical Abstract

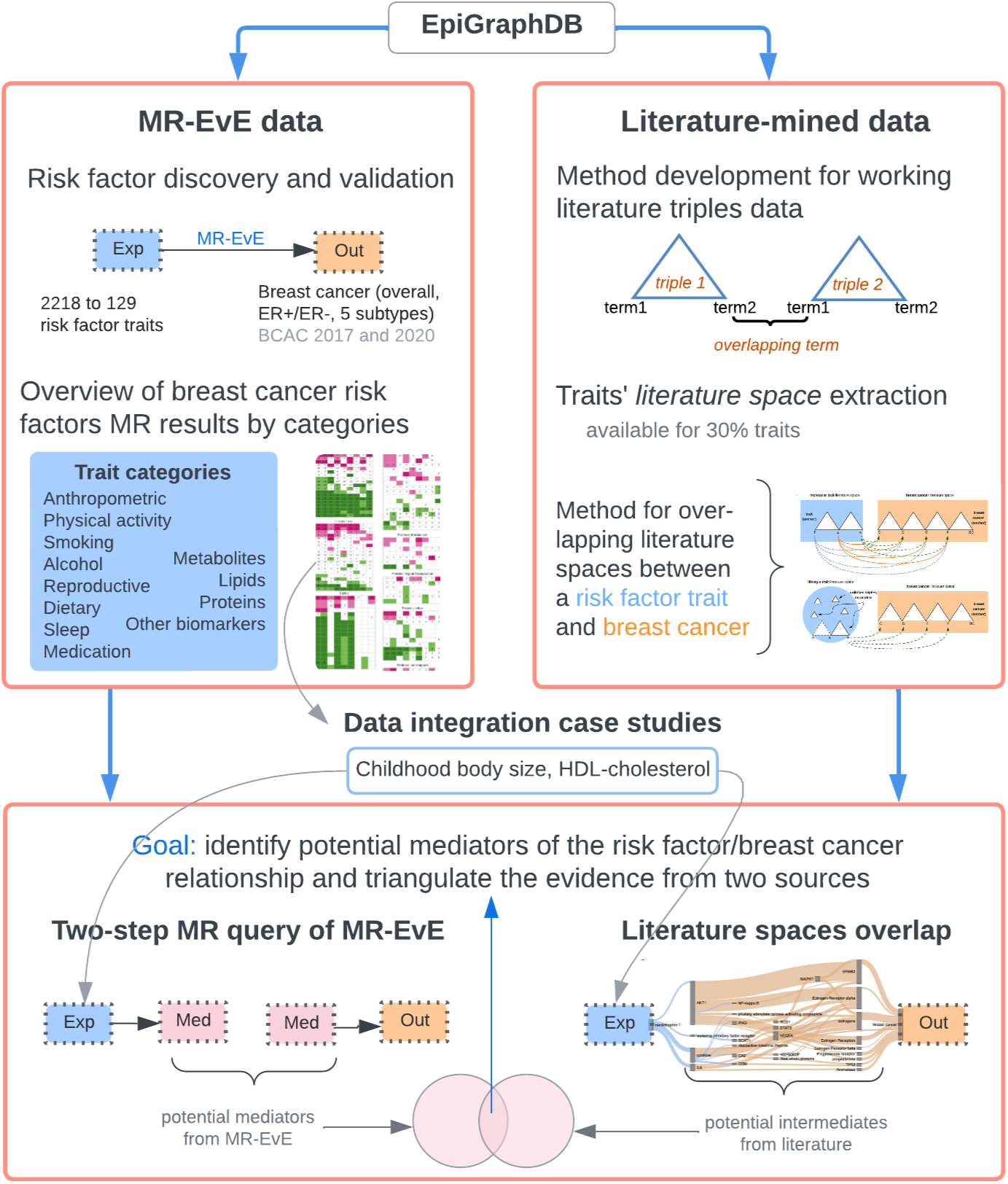

**Objective:** An increasing challenge in population health research is efficiently utilising the wealth of data available from multiple sources to investigate disease mechanisms and identify potential intervention targets. The use of biomedical data integration platforms can facilitate evidence triangulation from these different sources, improving confidence in causal relationships of interest. In this work, we aimed to integrate Mendelian randomization (MR) and literature-mined evidence from the EpiGraphDB biomedical knowledge graph to build a comprehensive overview of risk factors for developing breast cancer.

**Methods:** We utilised MR-EvE (“Everything-vs-Everything”) data to identify candidate risk factors for breast cancer and generate hypotheses for potential mediators of their effect. We also integrated this data with literature-mined relationships, which were extracted by overlapping literature spaces of risk factors and breast cancer. The literature-based discovery (LBD) results were followed up by validation with two-step MR to triangulate the findings from two data sources.

**Results:** We identified 129 novel and established lifestyle risk factors and molecular traits with evidence of an effect on breast cancer, and made the MR results available in an R/Shiny app (https://mvab.shinyapps.io/MR_heatmaps/). We developed an LBD approach for identifying potential mechanistic intermediates of identified risk factors. We present the results of MR and literature evidence integration for two case studies (childhood body size and HDL-cholesterol), demonstrating their complementary functionalities.

**Conclusion:** We demonstrate that MR-EvE data offers an efficient hypothesis-generating approach for identifying disease risk factors. Moreover, we show that integrating MR evidence with literature-mined data may be used to identify causal intermediates and uncover the mechanisms behind the disease.

## 1. Introduction

Triangulation, a process of integrating evidence from multiple methodologies, has become increasingly important in population health research [1, 2]. Different evidence sources are likely to have unique and unrelated sources of bias, so consistent evidence from such sources improves confidence in the causal relationship between a risk factor and outcomes of interest [3]. A potential obstacle to the widespread adaptation of triangulation in epidemiological research is the challenge of integrating and harmonising the large volume of data from different datasets. Multiple platforms [4, 5, 6, 7] have been developed to facilitate access to combined and curated biomedical datasets. Using these data integration platforms will facilitate the study of disease aetiology.

EpiGraphDB (https://epigraphdb.org) [4] is a biomedical knowledge graph that was designed to facilitate data mining of epidemiological relationships. The unique advantages of this platform are the availability of both Mendelian randomization (MR) [8, 9] and literature-mined relationships between molecular and lifestyle traits and disease outcomes, providing a valuable combination of data to systematically investigate causal relationships.

MR is an approach to causal inference that uses genetic variants as instrumentable variables (IVs) to infer whether a modifiable health exposure influences a disease outcome [8]. Studying disease causality with MR methods has been growing in popularity over the past decade and a half [9, 10], with the increasing availability of data from genome-wide association studies (GWAS) [11, 12] and well-developed analysis frameworks [13]. EpiGraphDB provides access to MR-EvE (Mendelian randomization “Everything-vs-Everything”) pairwise causal estimates [14] between thousands of traits from the OpenGWAS database (https://gwas.mrcieu.ac.uk) [11]. MR-EvE data can be used to explore potential causal links between thousands of traits for knowledge discovery or hypothesis generation.

Another aspect of knowledge discovery is extracting data from the biomedical literature, also known as literature-based discovery (LBD) [15]. In addition to MR-EvE, EpiGraphDB also contains literature-mined relationships extracted from publications in PubMed [16] and mapped to specific phenotypes in the MR-EvE data. LBD entails connecting information that is explicitly stated in multiple publications across different research disciplines aiming to deduce connections that have not been explicitly stated [17, 18], offering the potential to identify mechanistic pathways or mediators for causal relationships identified using MR-EvE.

In this work, we use EpiGraphDB to build a comprehensive picture of the aetiology of breast cancer by combining evidence from MR and literature-mined data. Breast cancer is a useful exemplar, as it is a heterogeneous disease with a complex aetiology, affected by both genetic and lifestyle factors [19, 20, 21, 22] that also has a large body of published research evidence, suitable for data mining. We seek to generate new mechanistic hypotheses for causal relationships using evidence integration to inform prevention and intervention strategies that could reduce disease burden.

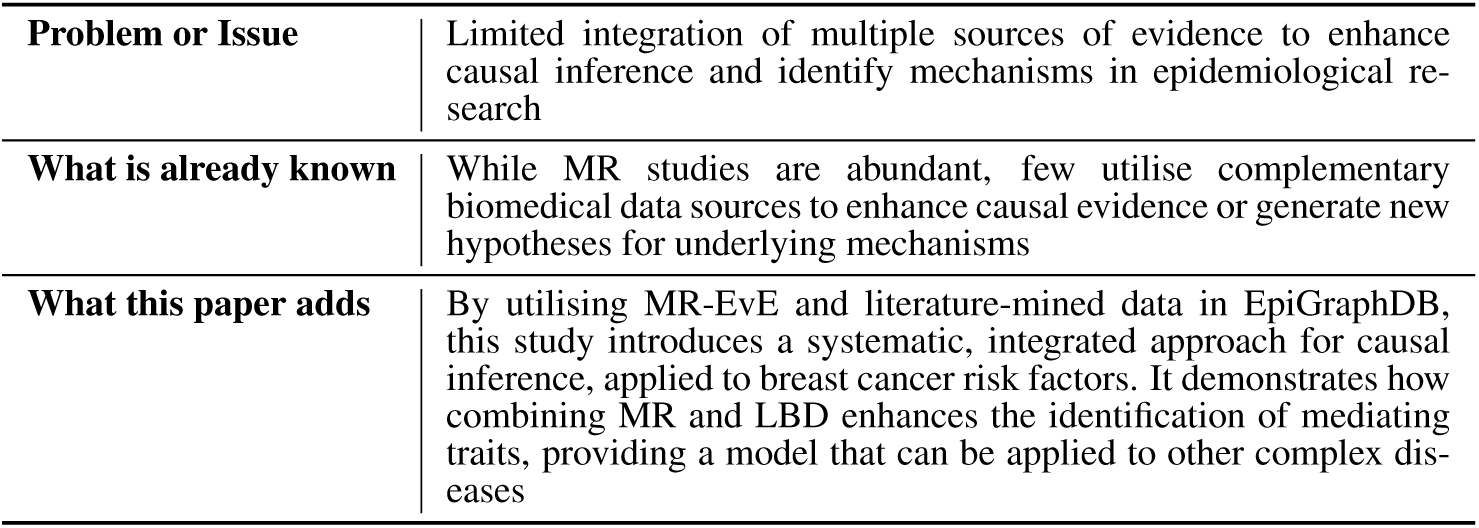
Statement of Significance.

## 2. Methods

### 2.1. Study Design

We present a summary of our EpiGraphDB data-focused study in Figure 1. First, we explored MR-EvE causal estimates between various exposure traits and breast cancer to generate a list of potential causal risk factors and biomarkers. Then, we used literature-mined relationships from EpiGraphDB to dissect the risk factor/breast cancer relationships. We designed an LBD method for overlapping literature spaces of traits to identify potential intermediates between individual traits and breast cancer.

**Figure 1:**
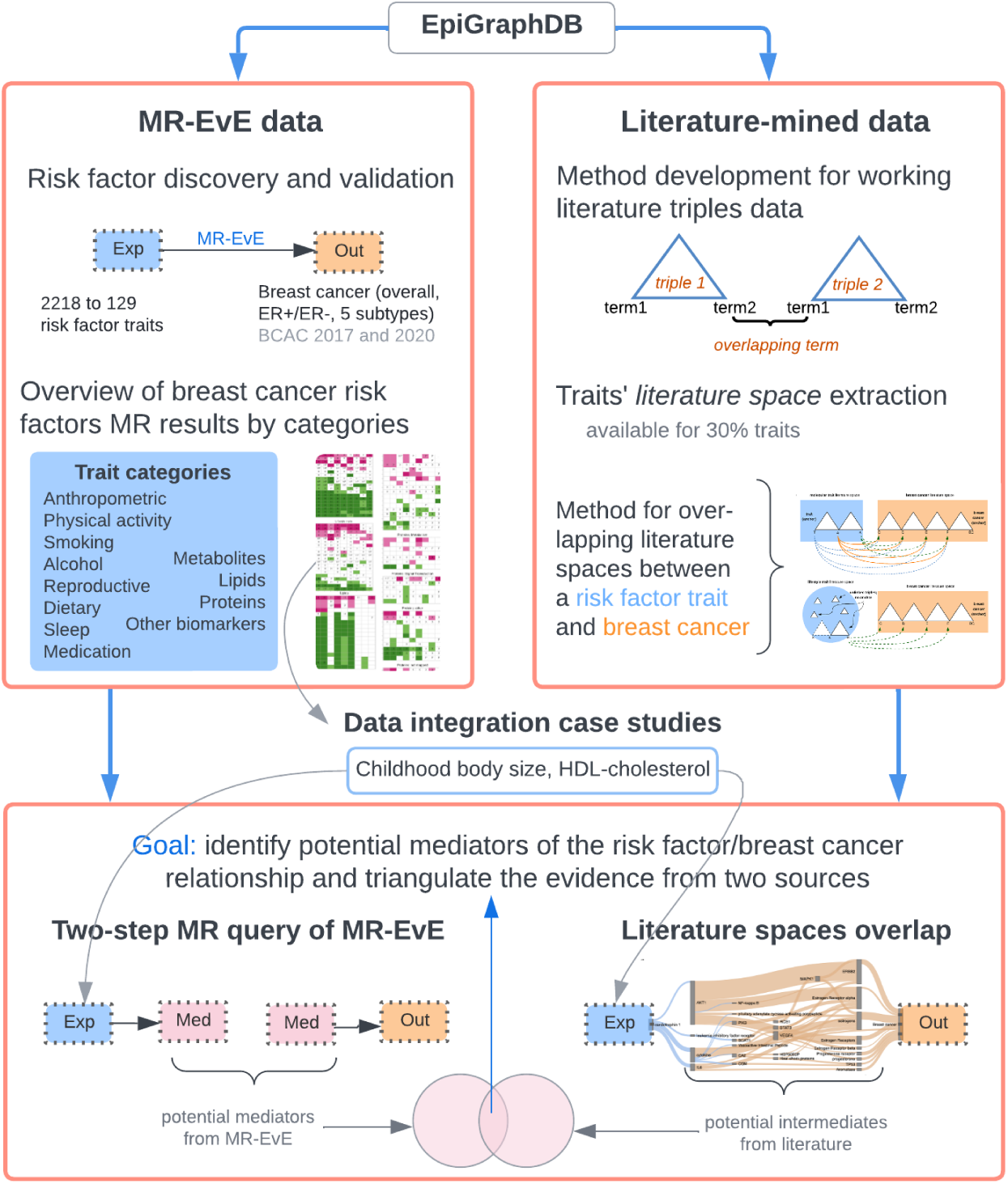
Study overview. Employing EpiGraphDB, MR-EvE data was used to identify breast cancer risk factors, and literature-mined data was used to design a literature-based discovery (LBD) method for identifying potential intermediates between risk factors and breast cancer. For two case studies, we present evidence integration between MR-EvE-guided and LBD-identified potential mediators. Abbreviations/definitions: MR-EvE – Mendelian Randomization “Everything-vs-Everything”; Exp – exposure; Med – mediator; Out – outcome; BCAC – Breast Cancer Association Consortium (refers to meta-analyses breast cancer cohort GWAS data); literature space – a collection of literature-mined relationships between pairs of biological terms related to a phenotype.

We demonstrate evidence integration between MR and LDB on two case study breast cancer risk factors: *childhood body size* and *HDL-cholesterol*. We queried MR-EvE data using a two-step MR approach [23, 24] to identify potential mediators (i.e. traits affected by each risk factor that in turn affect breast cancer risk). We also determined potential intermediates using LBD and evaluated them using MR and data from OpenGWAS. In this way, we triangulated evidence for the identified mediators from two independent data sources. Analysis code is available at https://github.com/mvab/epigraphdb-breast-cancer and https://github.com/mvab/epigraphdb_mr_literature_queries.

### 2.2. EpiGraphDB

EpiGraphDB (https://epigraphdb.org) is a graph database that integrates biomedical and epidemiological data, built to support data mining of risk factor/disease relationships. It contains trait relationships (observational and genetic), literature-mined relationships, biological pathways, protein-protein interactions (PPIs), drug–target relationships and other data sources [4]. EpiGraphDB is integrated with the OpenGWAS database data [11] (https://gwas.mrcieu.ac.uk), providing access to GWAS studies via the GWAS node of the graph. The number of GWAS studies in OpenGWAS at the time of writing is 50,040; of these, 34,494 are integrated within EpiGraphDB. EpiGraphDB (version 0.3.0) was accessed via R package *epigraphdb-r* (version 0.2.3).

### 2.3. Breast cancer data

EpiGraphDB provides access to multiple breast cancer GWAS datasets deposited in OpenGWAS. We used the most recent releases of the Breast Cancer Association Consortium (BCAC) (http://bcac.ccge.medschl.cam.ac.uk/) meta-analyses conducted in 2017 (BCAC 2017, N=228,951) [25] and 2020 (BCAC 2020, N=247,173) [26]. Since only BCAC 2017 is available in EpiGraphDB/OpenGWAS, this dataset was used for the MR-EvE risk factor discovery analysis. Both BCAC 2017 and 2020 were utilized for the validation stage. In addition to the overall sample, the datasets contain the ER+ and ER-sub-samples and five molecular subtypes (Supplementary Table 1). The study groups in the BCAC cohort do not include UK Biobank or other trait cohorts used in this study. The cohort includes European-ancestry individuals only.

### 2.4. Mendelian randomization and MR-EvE data

MR is an application of instrumental variable analysis where genetic variants are used as proxies (genetic instruments) to estimate the causal relationship between a modifiable health exposure and a disease outcome [8, 9]. MR-EvE (MR “Everything-vs-Everything”) data is stored as bidirectional relationships between GWAS traits in EpiGraphDB, available for 11,789 GWAS traits at the time of writing. The MR-EvE estimates were generated using the MR Mixture-of-Experts (MR-MoE) method by Hemani *et al* [14] before integration into EpiGraphDB. MR-MoE is a machine learning framework that automated the selection of instrument SNPs and the MR method for use in any specific causal analysis by predicting the model of pleiotropy and relating that prediction to the most appropriate model [14]. The method selects the MR ‘best estimate’ of causal relationships between the exposures and the outcome traits.

The interconnection of many traits in the MR-EvE data makes it a valuable resource for identifying potential mediators and confounders [27] for a causal relationship of interest (https://epigraphdb.org/confounder) [4]. In this work, we make use of this functionality to generate hypotheses for breast cancer risk factors and potential mediators.

### 2.5. Identifying breast cancer risk factor traits from MR-EvE

#### 2.5.1. Discovery

We queried EpiGraphDB to extract MR-EvE results for all available GWAS traits (N = 2218, Supplementary Table 2) with breast cancer as the outcome. Figure 2 summarises the discovery and validation stages of the analysis. The initial set of traits linked to breast cancer with any MR estimate was reduced to traits that passed False Discovery Rate (FDR), taking traits with the corrected p-value *<* 0.05 (N = 378) into the downstream analysis (Figure 2).

**Figure 2:**
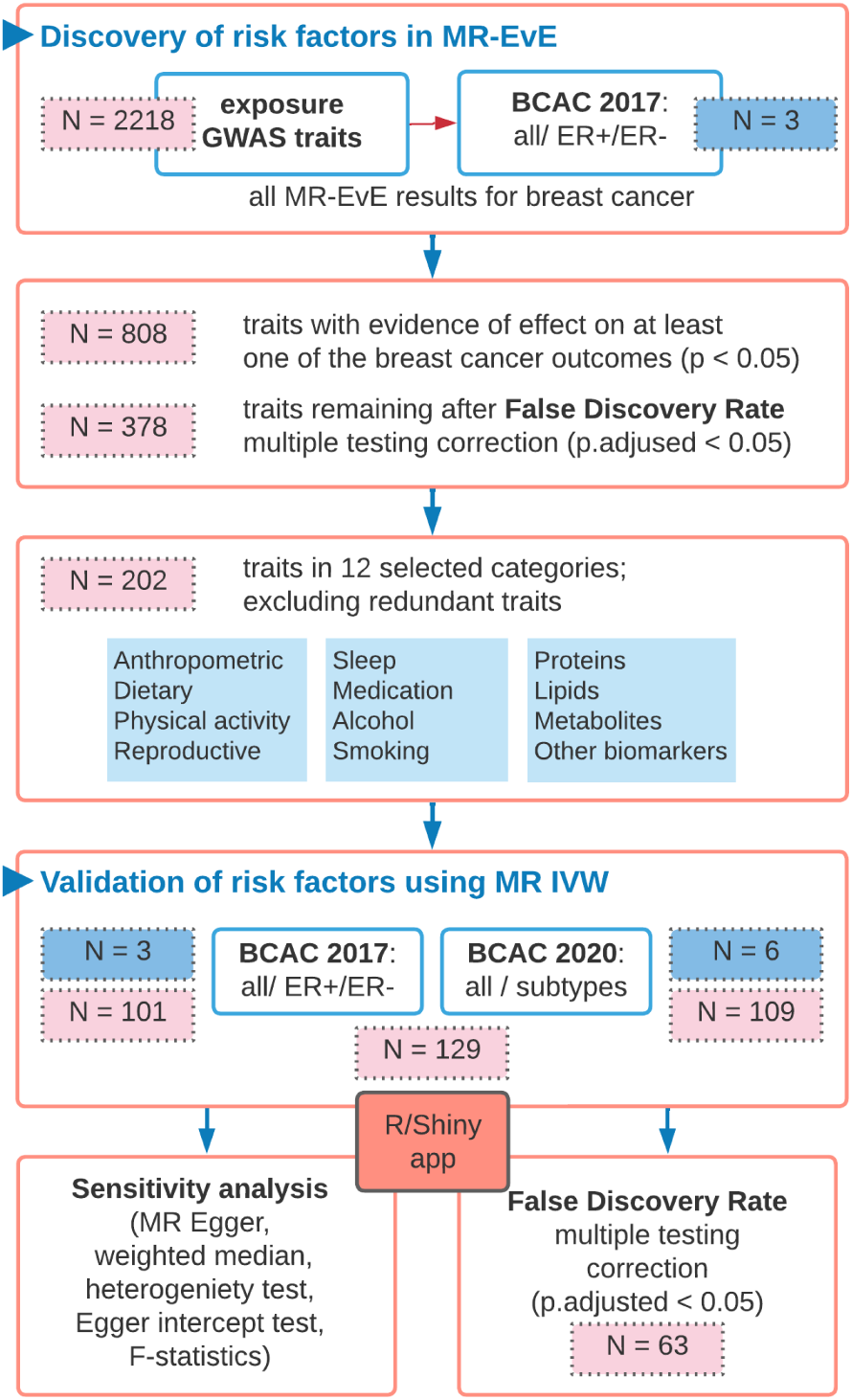
MR-EvE evidence processing workflow, including Discovery and Validation stages. The boxes show the number of exposures (pink) and breast cancer outcomes (blue) after/in each filtering step; MR-EvE results validation Shiny app: https://mvab.shinyapps.io/MR_heatmaps/. BCAC-Breast Cancer Association Consortium

The traits were grouped into 12 categories, which included widely-known cancer risk factor categories (anthropometric traits, reproductive traits, physical activity, smoking, alcohol), categories with a growing research interest (dietary intake traits, sleep traits, medication), and molecular traits (metabolites, lipids, proteins, and other biomarkers) (N = 202). The traits outside of these categories were grouped as ‘Other’ and excluded. The full data from the discovery stage is available in Supplementary Table 3.

#### 2.5.2. Validation

The results of the MR-EvE rapid screen of risk factors were validated by re-analysing the identified relationships using the robust practices in two-sample MR [28], because MR-EvE ‘best estimates’ selected by the MR-MoE method do not always correspond to the standard methods used in MR studies [14]. The validation results in this study are reported as per the STROBE-MR guidelines [29, 30].

For the majority of exposure traits, instruments were extracted using the genome-wide approach, i.e. SNPs with p-value *<* 5 *×* 10*^−^*^8^ and clumped with *r*2 *<* 0.001 using 1000 Genomes European LD reference panel. For protein traits, *cis*-region instruments were prioritised (within 1Mb on either side of the protein-encoding gene) [31]. The extracted *cis*-SNPs were filtered to p-value *<* 5 *×* 10*^−^*^8^ or a less stringent ‘local’ threshold of p-value *<* 0.05/nSNPs, where nSNPs is the number of SNPs in the *cis* region. The clumping was done using *r*2 *<* 0.01, or *r*2 *<* 0.001 if the higher threshold produced *≥* 7 SNPs. If no instruments were available under these criteria, genome-wide extraction of instruments was applied. The details of instrument extraction for protein traits are provided in Supplementary Tables 4 and 5.

The validation analysis was performed on breast cancer molecular subtypes data (BCAC 2020) in addition to the overall sample and ER+/ER-subtypes (BCAC 2017). The inverse-variance weighted (IVW) [32] approach was used as the main two-sample MR method. To correct for multiple testing in the validation step, the FDR cut-off of 0.05 for the corrected p-value was applied to identify results with more robust evidence of effect (Supplementary Table 4) (Figure 2).

To investigate potential violations of the MR assumptions and validate the robustness of MR results, additional analyses were performed. This included pleiotropy-robust sensitivity analyses such as MR-Egger [33] and weighted median [34]. The Egger intercept test [33] was used to test for directional horizontal pleiotropy, and Cochran’s Q statistic [35] was calculated to quantify heterogeneity among SNPs, which is indicative of potential pleiotropy. F-statistic was used to assess instrument strength [36] (Supplementary Tables 4 and 6). Sensitivity analysis results were not used for additional filtering of all identified risk factor traits to avoid information loss due to dichotomisation [37], but were taken into account when evaluating individual traits. Additionally, the sensitivity results were included in the MR results heatmap visualisation to provide context on the performance in sensitivity tests for all traits. All MR and sensitivity analyses were performed using the *TwoSampleMR* R package (version 0.5.6) [13].

### 2.6. Using literature-mined relationships in EpiGraphDB

EpiGraphDB contains literature-mined relationships between pairs of terms (also known as ‘literature triples’) derived from the published literature [38]. The underlying literature data comes from SemMedDB (Semantic MEDLINE Database) [16], a well-established repository of literature-mined semantic triples, i.e. ‘*subject term 1 – predicate – object term 2*’, mined from titles and abstracts of nearly 30 million biomedical articles in PubMed, using SemRep [39, 40] (further details are available in Appendix A). A subject/object term can be any biological entity (gene name, drug, phenotype, disease); a predicate is a verb that represents a relationship between the two terms (*affects, causes, inhibits, reduces, associated with*, etc.).

#### 2.6.1. Literature space

Literature triples of biomedical terms are mapped to specific GWAS traits in EpiGraphDB. In this work, we introduce the concept of a ‘*literature space*’. The literature space for a trait (e.g. breast cancer) comprises a set of semantic triples extracted from the published literature for this trait. Literature space can be considered an ‘automated literature overview’ of conceptual relationships between biological entities related to the search term, i.e. breast cancer. In the literature space, each relationship triple has a score that indicates how frequently it occurs in the published literature within a defined time frame (linked to PubMed IDs), demonstrating how well-established this relationship is.

#### 2.6.2. Literature spaces overlap method

In the field of LBD, the underlying principle of working with triple-based literature data is known as Swanson linking [41, 42]. In this approach, two separately published results (e.g. A-B and B-C relationships) are combined to identify evidence for the A-C relationship that is unknown or unexplored. The discovery can be open and closed: in open, A is given to identify Bs linked to Cs, generating hypotheses; in closed, A and C are fixed, and Bs that link them are identified, testing hypotheses or finding intermediates [43, 18] (details in Appendix A).

In this work, we present an extension to the simple triple linking method used in MELODI-Presto [38], which focused on the overlap of two triples (i.e. A-B and B-C only) (3a). Here, we combine multiple triples from two literature spaces (exposure trait and breast cancer) into a so-called *’literature overlap’*. We apply multi-step triple linking within and between the literature spaces to generate hypotheses for the potential mediators of the exposure trait’s effect on breast cancer. The literature data pre-processing steps and the method details are provided in Appendix A. The overview of our LBD method is presented in Figure 3, including closed (3b) and open (3c) discovery, and an example application (3d).

**Figure 3:**
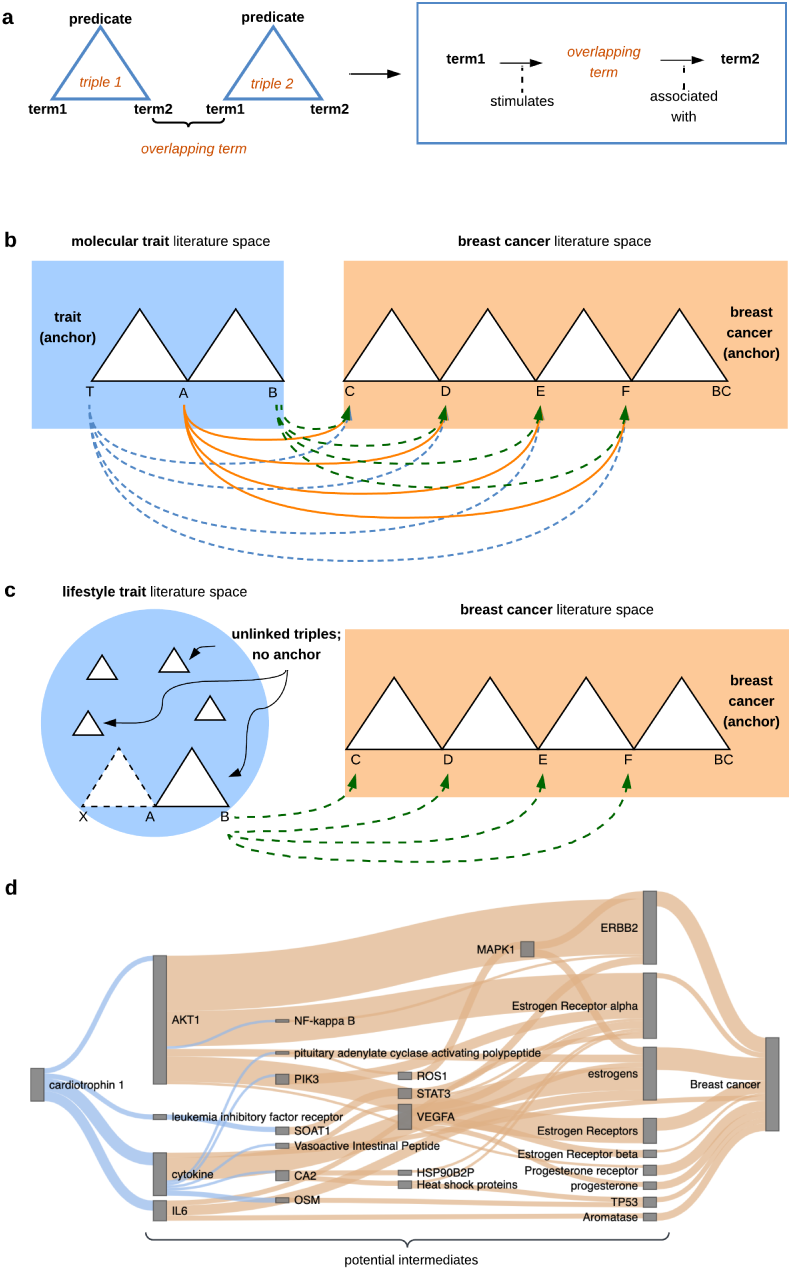
Schemas of the literature-based discovery method, demonstrating the approach behind literature spaces overlap. **(a)** A basic example of linking two triples via an overlapping term. **(b)** *Closed discovery*: overlap of two traits’ literature spaces (blue - risk factor, orange - breast cancer) by chaining triples of terms between them. The outer terms are restricted (‘i.e. ‘anchored’) to the term representing the trait (T: trait) and the term representing breast cancer outcome (BC: breast cancer). All intermediate terms are arbitrarily named A-F. The arrows represent all possible paths from T to BC, which are alternatives to the full path going through all intermediates. Closed discovery is applied to molecular traits. **(c)** *Open discovery*: Applied to non-molecular/lifestyle traits, where the trait is not available as a term and therefore cannot be anchored. The unlinked triples in the non-molecular trait space are first matched (via term B) to link with any triples in the breast cancer space. Then, the linked triples are connected to other triples in the non-molecular trait space (via term A), expanding the literature spaces overlap. **(d)** *Example of literature spaces overlap visualisation*: Sankey diagram showing how triples within and between a risk factor trait (here, cardiotrophin-1) and breast cancer spaces interconnect. The line width of each term relationship indicates how common it is in the literature (frequency score). Cardiotrophin-1 is presented here as an example. More examples, specifically the case study traits in this study, are available to view at https://mvab.shinyapps.io/literature_overlap_sankey/

### 2.7. MR and literature evidence integration

We integrated the evidence from MR-EvE and literature mining for potential mediators of two breast cancer risk factors that we selected as case studies: *childhood body size* and *HDL-cholesterol*.

#### 2.7.1. Potential mediators from MR-EvE data

We queried the MR-EvE data to identify potential mediators of each case study risk factor using the two-step MR framework [23, 24, 44]. The mediator search results were corrected for multiple testing using FDR at the individual case study level, for each step within two-step MR: *(step 1)* a risk factor trait effect on potential mediators, and *(step 2)* potential mediators’ effects on breast cancer outcomes (BCAC 2017 data only).

The mediators passing the FDR-adjusted p-value *<* 0.05 filtering in both steps and restricted to the earlier specified 12 trait categories were taken into the validation step. The validation for two-step MR results included the same analysis and sensitivity methods used for the main risk factors (2.5.2). The validated two-step MR estimates were also corrected for FDR.

#### 2.7.2. Potential mediators from the LBD literature overlap approach

The literature spaces for risk factor traits and breast cancer were constructed using the literature data linked to the following GWAS traits: breast cancer (combination of two traits: ‘*breast cancer*’ - *ieu-a-1126* and ‘*malignant neoplasm of the breast*’ - *finn-a-C3 BREAST*), HDL (*’HDL-cholesterol’* - *ukb-d-30760 irnt*), childhood body size (*’childhood obesity’*- *ieu-a-1096*) (Supplementary Tables 14-15).

In the literature overlap generated from triples in the risk factor trait and breast cancer literature spaces, all intermediate biological terms were evaluated as potential mediators. Each term was searched in OpenGWAS for representative GWAS data to be used in MR analysis, and genetic instruments for the identified traits were extracted as previously described (2.5.2).

We performed MR bidirectionally between the risk factor trait and each intermediate to identify the direction of effect, as it may not be correct in literature-mined relationships. Then, the intermediate traits were evaluated for an effect on breast cancer. The results were validated using the analysis and sensitivity methods described previously (2.5.2), and adjusted using FDR.

The evidence in both steps of the two-step MR analysis provided suggestive evidence of potential mediation between the risk factor and breast cancer via a trait identified by the LBD method.

## 3. Results

### 3.1. Breast cancer risk factors discovered from MR-EvE

We identified 129 traits across 12 categories that showed evidence of an effect on breast cancer risk from MR-EvE data (Figure 2). The discovery effect estimates were validated using the IVW MR analysis with sensitivity analyses, using 9 breast cancer outcome GWAS from two releases of BCAC data. The instruments were extracted as described in 2.5.2. To correct for multiple testing of many exposures and outcomes in the validation step, the FDR correction was applied. The validation results and sensitivity analyses are available in Supplementary Tables 4 and 6.

The MR results for the final set of traits (129 traits or 63 after FDR correction) are presented in Figure 4. The figure also provides an overview of sensitivity analysis results: potential pleiotropy “P”, heterogeneity “H”, and “X” for exposures with a low number of instruments (*≤* 2) where most sensitivity tests were not possible. The full results (202 traits) are available in the R/Shiny app (https://mvab.shinyapps.io/MR_heatmaps/) and Supplementary Table 4. The abbreviations for traits/biomarkers used throughout the section are defined in Supplementary Table 7. In the sections below, the results of several trait categories are summarised.

**Figure 4:**
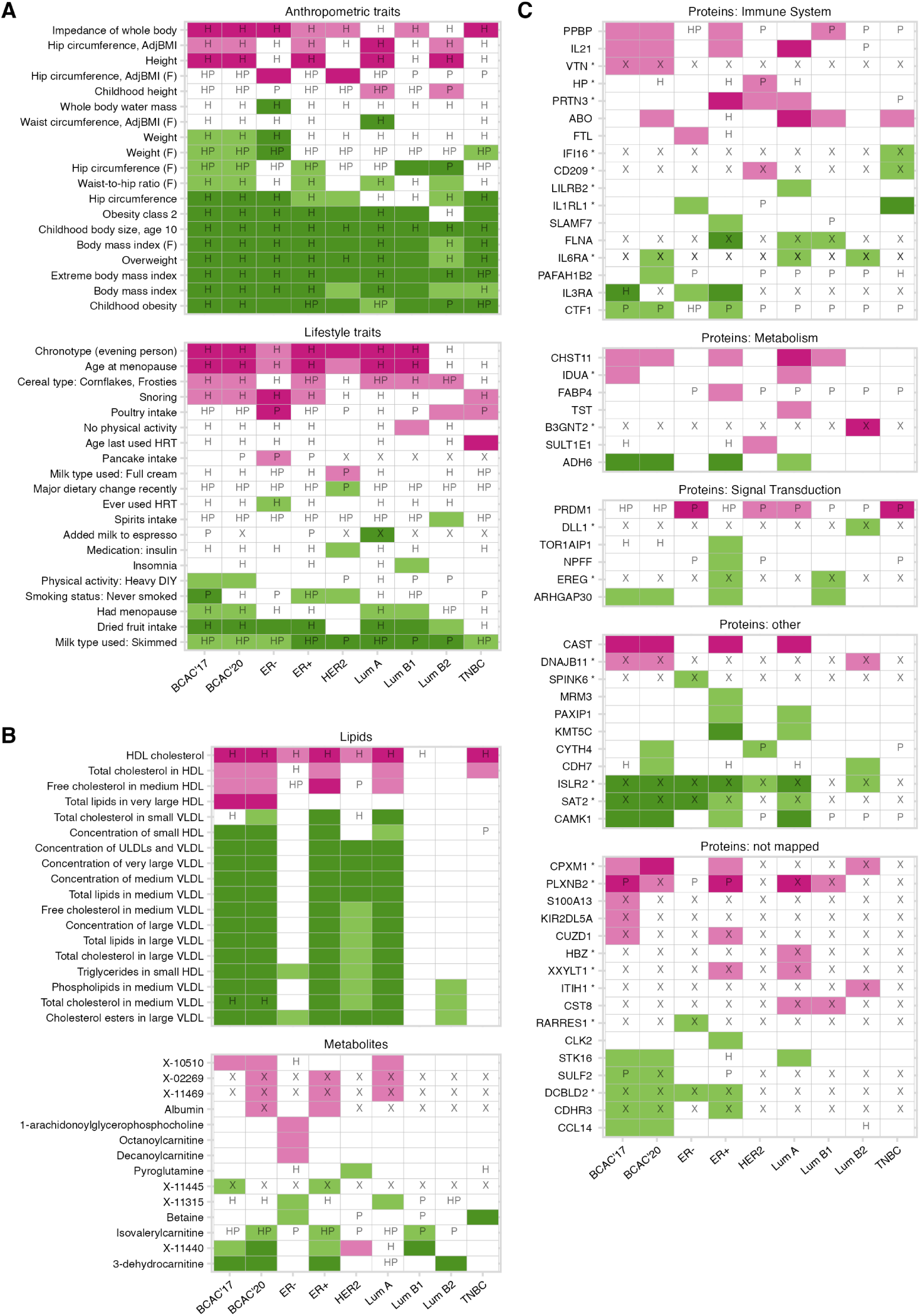
Mendelian randomization effect direction heatmaps for (A) lifestyle and anthropometric traits, (B) metabolites and lipids, (C) proteins (N=129) as exposures, and breast cancer (BCAC 2017 and 2020 overall samples and subtypes) as the outcomes. The presented traits have evidence of an effect on at 1le3ast one outcome, but the full set of traits analysed in the validation step (N=202) is available in the interactive version of the plot in the app: https://mvab.shinyapps.io/MR_heatmaps/ and in Supplementary Table 4. The proportions of the total size of each group displayed here are: Anthropometric – 95%, Lifestyle – 57%, Lipds −100%, Metabolites – 68%, Proteins – 54%. The colours represent effect direction: pink – positive (causal), green – negative (protective), white – null effect (based on 95% CIs of odds of breast cancer per standard deviation increase in the exposure). The darker colour shade indicates the results that passed the FDR correction. The letters summarise arbitrarily dichotomised sensitivity analysis results: “P” - potential pleiotropy (*abs*(egger intercept)*>*0.05); “H” - heterogeneity (Q-stat p-value *<* 0.05); “X” - low number of instruments (*≤* 2) where sensitivity analyses were not possible; an empty cell indicates that no H/P/X issues were identified. For proteins, where possible, *cis*-only instruments were used (marked with an asterisk *). The proteins were grouped by major pathways defined in the Reactome database [45] for display purposes. Proteins are displayed by the encoding gene name (defined in Supplementary Table 7). The exposure traits are from mixed-sex samples unless otherwise specified (F: female-only) or are female-specific reproductive traits. The details on breast cancer outcome samples are provided in Supplementary Table 1.

#### 3.1.1. Non-molecular traits

The majority of anthropometric traits (e.g. *body mass index, obesity, waist-to-hip ratio, childhood body size*) (Figure 4a) have consistent inverse (risk-decreasing) effects across most breast cancer subtypes. Interestingly, *impedance of whole body* has a positive (risk-increasing) effect on most outcomes, potentially capturing a different aspect of adiposity. The traits in this group show evidence of high instrument heterogeneity (‘H’ in Figure 4a) based on the arbitrary threshold used (Q-stat p-value *<* 0.05). This indicates the complexity of these phenotypes, suggesting that multiple underlying mechanisms are likely involved. Evaluating multiple related/correlated phenotypes is acceptable, as none of them are perfect measures of adiposity, and they likely proxy adiposity in different ways, e.g. BMI (subcutaneous fat) vs waist-hip ratio (central adiposity).

Among physical activity traits (UK Biobank traits based on a survey question *”Types of physical activity in last 4 weeks”*), *‘Heavy DIY’* shows evidence of a protective effect in overall samples (with little indication for heterogeneity and pleiotropy). Among dietary traits, there is a protective effect from *‘dried fruit intake’* on outcomes with larger sample sizes. The intake of *processed/sugary breakfast cereals (i.e. Frosties, Cornflakes)* appears to have a risk-increasing effect, but the instruments here are also likely highly pleiotropic as is commonly seen for dietary intake traits. The important considerations for dietary and physical activity exposures in MR analyses are addressed in Appendix C.

The *‘never smoked’* phenotype appears to confer a decreased risk in selected breast cancer subtypes. *‘Chronotype (evening preference)’* shows evidence of increasing the risk in outcomes with larger sample sizes. *‘Snoring’* also has a causal effect on some outcomes. Higher *age at menopause* has a risk-increasing effect on most outcomes, while ‘*Had menopause*’ phenotype (potentially capturing earlier menopause) is protective.

#### 3.1.2. Molecular traits

The lipid traits have a distinct effect direction pattern (Figure 4b): a consistently positive (causal) effect from HDL particle measures and a negative (protective) effect from VLDL. For all, there is limited evidence of effect on ER-breast cancer and several molecular subtypes. It should be noted that LDL measures were also available in the MR-EvE discovery data, (i.e. 2218 traits), but did not pass filtering thresholds in the validation step due to weak evidence of effect (data available in Supplementary Table 3). Among other metabolites (Figure 4b), 3-dehydrocarnitine, betaine, and X-11440 show the strongest evidence of a negative effect on breast cancer risk.

The proteins with the most robust evidence (FDR-adjusted p-value *<* 0.05 and based on *cis*-instruments only) are IL1RL1, ISLR2, SAT2 (negative effect) and PRTN3, B3GNT2, CPXM1, PLXNB2 (positive effect). The proteins that did not pass FDR correction, but are also based on *cis*-only instruments and have effect on more than one outcome, include IL6R, EREG, DCBLD2 (negative effect) and VTN, IDUA, DNAJB11, XXYLT1 (positive effect).

Among the proteins for which genome-wide instruments were used (which may be picking up horizontal pleiotropy), the most consistent evidence of effect was observed for FLNA, ADH6, CAMK1 (negative effect) and IL21, CAST (positive effect) with little heterogeneity and pleiotropy detected (‘P’ and ‘H’ not present in Figure 4c).

### 3.2. Evidence integration case studies

This section presents two exposure traits from the discovery of the MR-EvE risk factor, *childhood body size* and *HDL cholesterol*, as case studies. Although both have previously been investigated in relation to breast cancer, the underlying mechanisms remain unclear. We used MR-EvE data and data mined from literature to generate hypotheses for potential mediators and cross-validate evidence from both sources. Since these traits have opposite effects on breast cancer risk and represent both molecular and non-molecular risk factors, they provide a robust framework for demonstrating open and closed literature-based discovery, supported by large literature spaces (Appendix A).

#### 3.2.1. Case study 1: Childhood body size

Childhood adiposity has been shown to have a protective effect on breast cancer risk in both observational [46, 47] and MR studies [48, 44], but the mechanism and mediators of this effect remain unclear. MR analysis shows strong evidence of an inverse effect from childhood body size (OpenGWAS ID: *ukb-b-4650*, female-only sample) on all breast cancer subtypes (Figure 4) (e.g. OR 0.60 [95% CI 0.53: 0.69] for BCAC 2017 overall sample).

##### MR-EvE mediators

In the initial MR-EvE scan, 125 putative mediators of the childhood body size effect on breast cancer were identified. Among these, several were considered as potential mediators in earlier work [44]: IGF-1, age at menarche, age at menopause, age at last birth, glucose, glycated haemoglobin (Supplementary Table 8). Following two-step MR results validation, 71 traits had evidence of effect in both steps at nominal significance and 54 traits at the FDR-corrected level (Supplementary Tables 9 and 10). Among these, a large proportion were adult anthropometric traits (N=30). Since adult adiposity has been shown to be unlikely to mediate the childhood adiposity effect [48, 49], these traits were not reviewed further. Lipid traits (N=20) were also not considered further as they are unlikely to mediate substantial effect individually. The remaining candidate mediators traits were: *exercise to keep fit* (from UK Biobank physical activity questionnaire), *sleep duration, skimmed milk intake, and Apolipoprotein A (ApoA)* (Table 1).

**Table 1:**
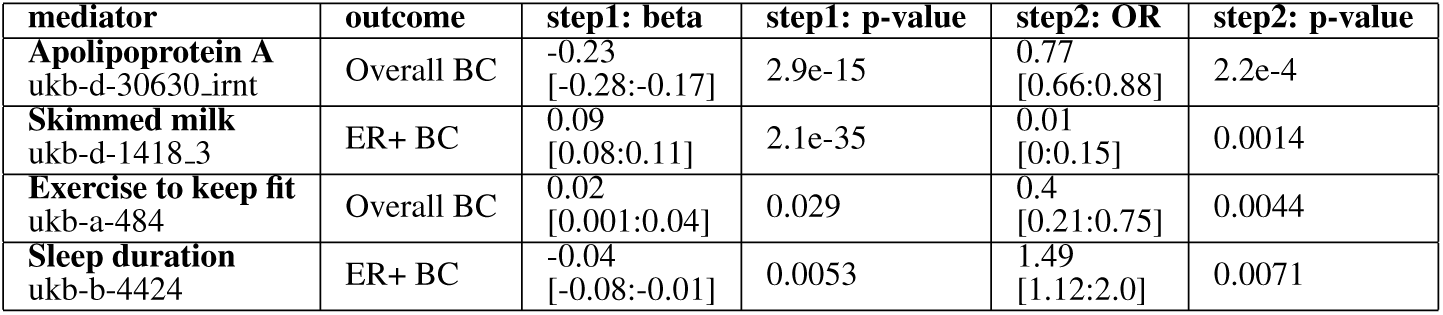
Two step-MR results for potential mediators of childhood body size/breast cancer relationship, identified in a hypothesis-free way via mining MR-EvE. The results in the table are from the MR IVW validation analysis, showing beta [95%CI] for *step 1* (the effect of childhood body size on mediator, measured as SD change in mediator per body size category change) and OR [95%CI] for *step 2* (the odds of breast cancer per SD higher mediator). The table only includes the results that passed FDR in both steps. The table does not include anthropometric and lipid traits (the full results are available in Supplementary Table 9).

Higher childhood body size shows some evidence of an increasing effect on the amount of *exercise to keep fit* in adulthood (effect size: 0.02 [95% CI 0.001: 0.04], step 1), which in turn has a protective effect on breast cancer risk (OR: 0.4 [95% CI 0.21: 0.75], step 2) (in agreement with published MR study [50] and results in 3.1.1). The relationship between childhood adiposity and the amount of exercise in adulthood has not been explored with MR designs before, so this finding warrants further follow-up.

Higher childhood body size has a negative effect on *sleep duration* in adulthood (effect size −0.04 [95% CI −0.08: −0.01], step 1), while longer sleeping time increases breast cancer risk (stronger evidence for ER+, OR 1.49 [95% CI 1.12: 2.0], step 2), in agreement with published work [51].

##### Literature mediators

Next, we reviewed literature-mined terms that connect childhood adiposity to breast cancer. The Sankey diagram in Supplementary Figure B.6 shows the literature overlap between the two traits (Supplementary Table 17), performed using the open discovery approach (Figure 3c, 2.6.2) (view interactively at https://mvab.shinyapps.io/literature_overlap_sankey/).

The intermediate terms of the literature overlap include traits considered in earlier hypothesis-driven work [44] as potential mediators: IGF-1, testosterone, SHBG, insulin, and oestradiol. IGF-1 was also found using MR-EvE, but did not pass the FDR correction (mixed-sex sample data). The overlap contains many other literature-mined terms that make interesting mediation hypotheses (e.g. leptin, prolactin, adiponectin).

Next, to assess the identified intermediate terms as potential mediators using MR analysis, we searched OpenGWAS for suitable GWAS data to represent them. Of the 112 potential intermediate traits identified, data was available for 49, with 38 traits having at least one SNP instrument (Supplementary Table 18).

We carried out MR bidirectionally between childhood body size (*ukb-b-4650*) and 38 potential intermediate traits (Supplementary Tables 19 and 20), followed by assessing their effect on breast cancer (Supplementary Table 21). A subset of combined results from the three MR analyses are presented in Table 2.

**Table 2:**
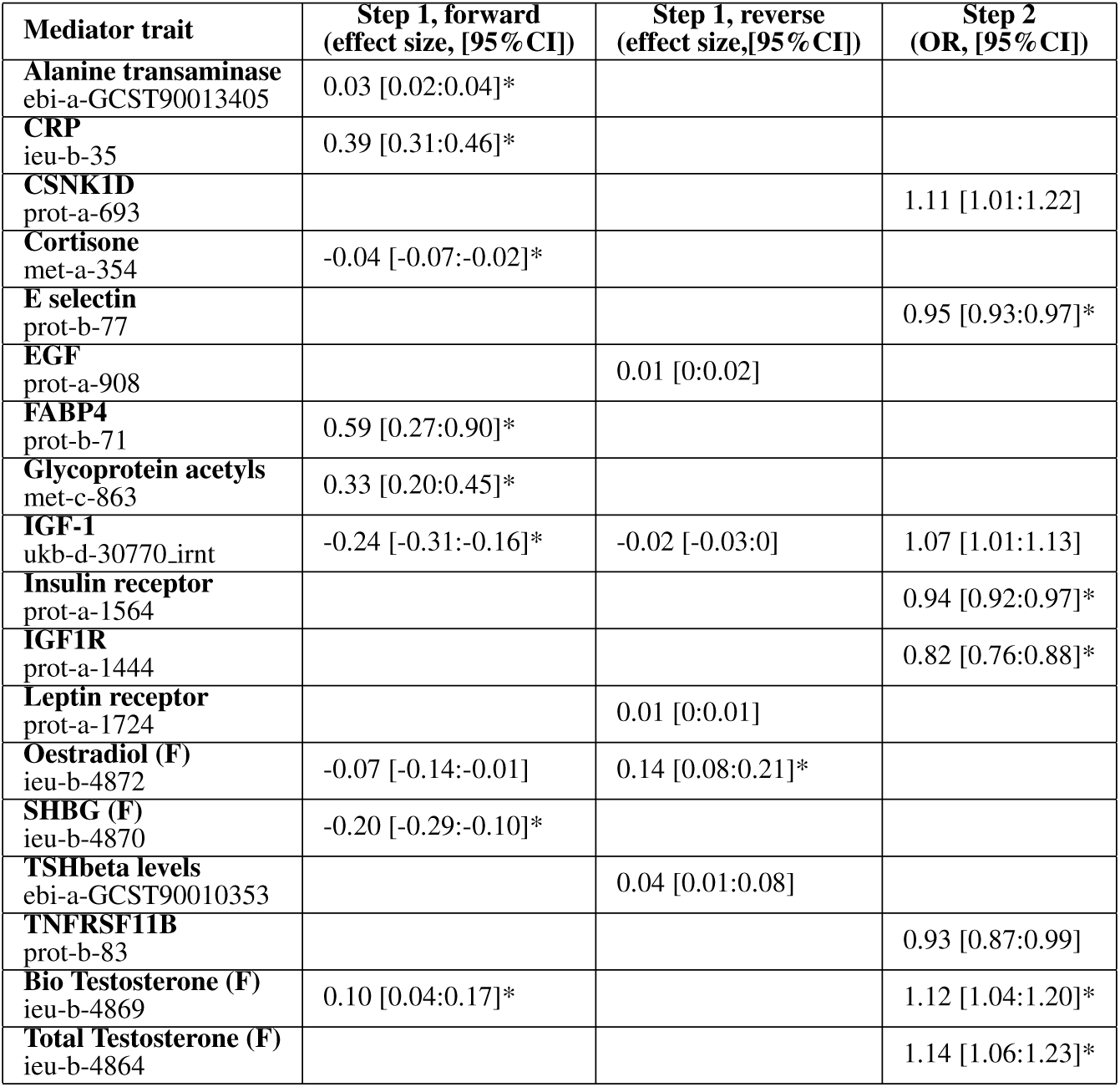
Subset of MR results (bidirectional and two-step) for potential mediators of childhood body size/breast cancer relationship, identified using LBD. The table combines the results from three MR analyses: *step 1 forward* (childhood body size *→*mediator trait), *step 1 reverse* (mediator trait *→* childhood body size), and *step 2* (mediator trait *→* overall breast cancer). ‘*step 1 forward*’ and ‘*step 1 reverse*’ corresponded to bidirectional MR analysis; ‘*step 1 forward*’ and ‘*step 2*’ together correspond to two-step MR study design. The table only shows the results that pass the threshold of p.value *<* 0.05; the results that pass FDR are marked with an asterisk*. The full results are available in Supplementary Tables 19, 20, 21. *Abbreviations*: CRP - C-Reactive protein, TSHbeta - Thyrotropin subunit beta, TNFRSF11b – tumour necrosis factor receptor superfamily member, CSNK1D - Casein kinase I isoform delta, EGF – epidermal growth factor, FABP4 - fatty acid binding protein 4; (F) - female-only data

The criteria for a trait to have suggestive evidence of being a potential mediator (i.e. evidence of effect in *step1 forward* and *step2* (i.e. two-step MR), and preferably no effect in *step1 reverse*) was met by two traits: IGF-1 and bioavailable testosterone. These two traits have been previously evaluated in [44] two-step MR analysis, with equivalent results and sensitivity analyses.

#### 3.2.2. Case study 2: HDL-cholesterol

HDL-cholesterol (HDL-C) has been shown to have a risk-increasing effect on breast cancer in several MR studies [52, 53, 54], which opposes the finding from a large meta-analysis of prospective studies that showed a modestly inverse association [55]. This makes HDL-C an interesting case study for hypothesis-free mediators search, as this may help to dissect the mechanism linking it to breast cancer.

The HDL-C trait in MR-EvE is based on mixed-sex sample data from UK Biobank (OpenGWAS ID: *ukb-d-30760 irnt*); the evidence of effect from IVW for BCAC 2017 overall sample is OR 1.09 [95% 1.04: 1.13] (no effect on certain subtypes) (Figure 4b), which is in agreement with previous studies and consistent between sensitivity analysis MR methods (Supplementary Tables 4 and 6).

##### MR-EvE mediators

In the initial MR-EvE scan, 102 potential mediators of HDL-C’s effect on breast cancer were identified, primarily lipid traits, proteins, and anthropometric traits (Supplementary Table 11). After two-step MR validation, 60 traits showed nominal significance, and 44 remained significant after FDR correction (Supplementary Tables 12 and 13). Of these potential mediators, 36 were lipid traits, which we did not evaluate further due to their high correlation with HDL-C and, therefore, the need for multivariable MR (MVMR) analysis. The identified mediators also included several anthropometric traits, which may have a role in the effect of HDL-C on breast cancer risk, but also would require further investigation using MVMR for traits capturing different adiposity components (e.g., waist-hip ratio, BMI).

HDL-C was found to have an increasing effect on two protein traits, *ApoA* (effect size: 0.87 [95% 0.84: 0.90]) and *cathepsin F* (CTSF) (0.14 [95% 0.03: 0.24]) (Table 1. The relationship and interplay between HDL-C, ApoA, and cathepsins have been investigated before [56, 57]. ApoA is the major protein component of HDL, which has an inverse effect on breast cancer in MR (OR: 0.77 [0.66:0.88], using *cis*-instruments only), also previously studied in connection to breast cancer [58]. CTSF has a positive effect on breast cancer risk (OR: 1.14 [95% 1.08: 1.21], single *cis*-instrument). CTSF belongs to a class of evolutionally conserved cysteine proteases involved in various biological processes that have been linked to disease, including cancer progression [59, 60]. Cathepsins (F, S, K; each to various degree) degrade lipid-free molecules of ApoA, leading to a complete loss of the ApoA’s function as a cholesterol acceptor, stopping its anti-inflammatory action [56], which may be related to CTSF’s role in disease. Further investigation into the mechanism linking HDL-C, ApoA, and CTSF (and potentially other cysteine proteases), and breast cancer is needed.

##### Literature mediators

Next, we reviewed literature-mined data connecting HDL and breast cancer to look for potential intermediates. Supplementary Figure B.7 shows the Sankey diagram of literature overlap between the traits (Supplementary Table 22), performed using the closed discovery approach (Figure 3b, 2.6.2) (view interactively at https://mvab.shinyapps.io/literature_overlap_sankey/).

The intermediate terms of this literature overlap include 120 unique (mostly molecular) traits. Many of them are lipids-related traits (e.g., LDL, VLDL, ApoA, ApoB, Lipase); others include interesting hypotheses such as testosterone, IGF-1, insulin, CRP, and cathepsin D. Suitable GWAS datasets for MR validation were found for 50 of the 120 traits (Supplementary Table 23).

We carried out MR bidirectionally between HDL-C (*ukb-d-30760 irnt*) and 42 potential intermediate traits (*>*=1 SNP) (Supplementary Tables 24 and 25), followed by assessing their effect on breast cancer (Supplementary Table 26). A subset of combined results from these three MR analyses is presented in Table 4. From these results, the traits that fit the criteria for suggestive evidence of mediation are ApoA and testosterone. ApoA has been identified as a potential mediator using MR-EvE as well. HDL-C has opposite effects on two types of testosterone: negative on bioavailable and positive on total (Table 4), potentially indicating different interaction mechanisms. Both testosterone measures have a risk-increasing effect on breast cancer (previously reported in [61, 44]). In the literature overlap, HDL is linked to testosterone via ApoE, HSD11B1, lipoproteins, and directly. In turn, testosterone is linked to breast cancer directly or via estrogen/progestin (Figure B.7).

Finally, cathepsin D (CTSD) has been identified as a potential intermediate in the literature overlap (via CETP/endopeptidases and lipoprotein/cyclosporine, Figure B.7). MR results show some evidence of HDL-C effect on CTSD (effect size −0.12 [95% CI −0.23:-0.003], but little evidence of CTSD effect on breast cancer (OR: 1.02 [95% CI 0.99:1.05], three *cis*-SNPs) (Table 4, Supplementary Table 26), suggesting an unlikely role of CTSD as a mediator. However, the discovery of a CTSD in literature overlap complements the finding of CTSDF in the MR-EvE data (Table 3), indicating that the cathepsins may have a role in the relationship of HDL-C and breast cancer, and the entire class of these enzymes should be explored in future work.

**Table 3:**
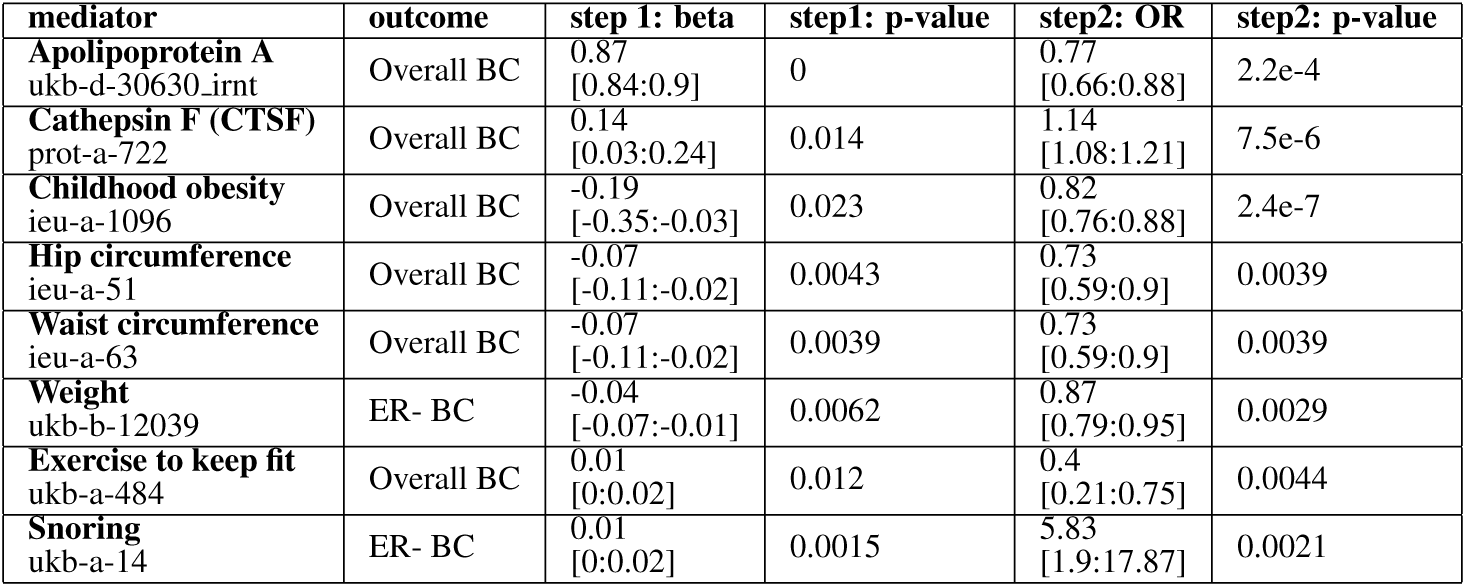
Two step-MR results for potential mediators of HDL-cholesterol/breast cancer relationship, identified in a hypothesis-free way via mining MR-EvE. The results in the table are from the MR IVW validation analysis, showing beta [95%CI] for *step 1* (the effect of HDL-C on mediator, measured as SD change in mediator per SD higher HDL-C) and OR [95%CI] for *step 2* (the odds of breast cancer per SD higher HDL-C). The table only includes the results that passed FDR in both steps. The table does not include lipid traits (the full results are available in Supplementary Table 12).

**Table 4:**
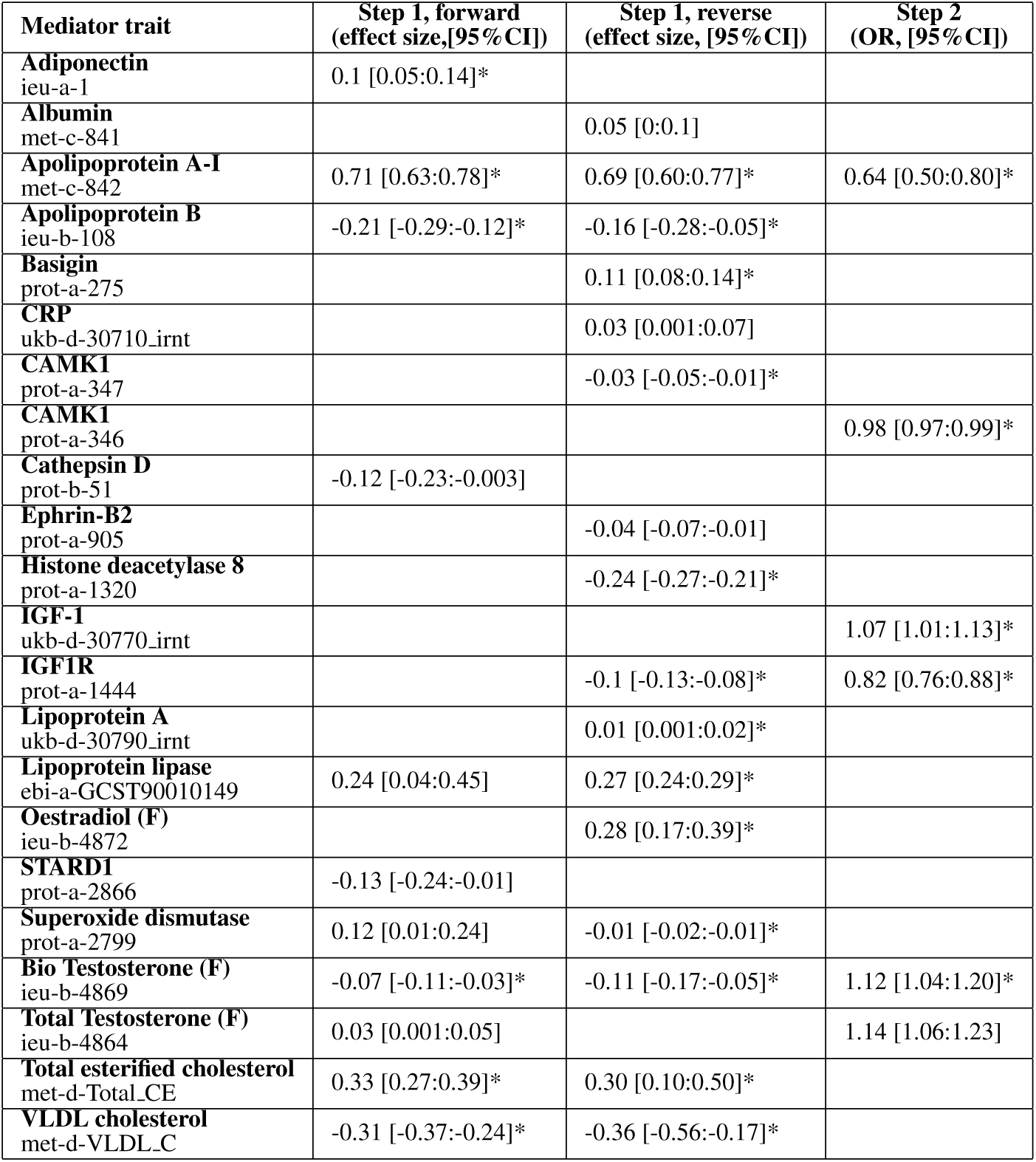
Subset of MR results (bidirectional and two-step) for potential mediators of HDL-C/breast cancer relationship, identified using LBD. The table combines the results from three MR analyses: *step 1 forward* (HDL-C *→*mediator trait), *step 1 reverse* (mediator trait *→* HDL-C), and *step 2* (mediator trait *→* overall breast cancer). ‘*step 1 forward*’ and ‘*step 1 reverse*’ corresponded to bidirectional MR analysis; ‘*step 1 forward*’ and ‘*step 2*’ together correspond to two-step MR study design. The table only shows the results that pass the threshold of p.value *<* 0.05; the results that pass FDR are marked with an asterisk*. The full results are available in Supplementary Tables 24,25,26. *Abbreviations:* CRP - C-Reactive protein, CAMK1 - Calcium/calmodulin-dependent protein kinase type 1, STARD1-Steroidogenic acute regulatory protein; (F) - female-only data

## 4. Discussion

In this study, we used the EpiGraphDB knowledge graph to systematically identify breast cancer risk factors. Additionally, we integrated literature-mined and genetic data to determine potential mediators of the identified causal relationships between these risk factors and breast cancer, which we presented via two case studies.

Through mining MR-EvE data in EpiGraphDB we identified 2218 traits affecting breast cancer risk, which were narrowed down to 129 traits with reliable causal evidence following rigorous validation (Figure 4, https://mvab.shinyapps.io/MR_heatmaps/). The identified risk/protective factors included both molecular biomarkers and non-molecular traits. Many of these traits have been reported before, e.g. anthropometric measures [48, 62, 63], reproductive traits [44], sleep traits [51, 64], height [65], smoking behaviour [66, 67], milk intake [68, 69], physical activity [70, 50, 71], lipids [52, 53, 54], metabolites [72], groups of proteins, e.g. adipokines [73], cytokines [74], inflammatory markers [75], metalloproteinases [76] and other circulating proteins [77, 78]; additionally, several studies evaluated multiple risk factor groups [44, 79, 80, 71]. Several molecular traits that have not previously been reported in MR studies include betaine, SAT2, PRTN3, PLXNB2, DCBLD2, IDUA. Appendix C provides an extended discussion of MR results of dietary and physical traits.

We presented two case study traits (*childhood body size* and *HDL-C)*, for which we identified potential mediators from MR-EvE data and literature data using our LBD method, aiming to triangulate the evidence from both sources. Two-step MR and sensitivity analyses were used to provide suggestive evidence of mediation through the identified traits. In the *childhood body size* case study, MR-EvE and literature-based mediator searches detected several overlapping traits, many of which have been considered as potential mediators in an earlier hypothesis-driven study [44] (e.g. IGF-1 and bioavailable testosterone), offering proof-of-concept evidence for this approach, in addition to identifying several traits not considered previously (e.g. physical activity and sleep duration). For *HDL-C*, the literature overlap method identified several intermediates that also appeared in the MR-EvE data (e.g. other lipid traits, ApoA, a cathepsin). HDL-C was also found to differentially affect total and bioavailable testosterone, which needs further investigation. Additionally, the entire class of cathepsin enzymes needs to be assessed for their mediating role in future work.

In our study, we took advantage of the pre-generated MR-EvE estimates in EpiGraphDB through the R package (*epigraphdb-r*) [4], and demonstrated how it enables fast hypothesis generation and prioritisation of candidate traits for more targeted investigation. Originally, MR-EvE was developed to provide a causal map of the human phenome, useful for supporting or rejecting specific hypotheses [14]. However, for most traits, patterns of horizontal pleiotropy and issues with invalid instruments were detected [14]. This highlighted the need for rigorous followup, including sensitivity analyses, biologically informed instrument selection, and triangulation with other sources of evidence to validate the associations [14, 81]. Hence, we validated our discoveries from MR-EvE using best practices in MR, accompanied by sensitivity analyses, *cis*-instrument extraction for protein traits, and also correction for multiple testing.

The MR-EvE data comes with several other limitations. Firstly, it is restricted to the traits available in OpenGWAS as of 2021, which were analysed in the everything-vs-everything way and subsequently added to EpiGraphDB. Secondly, a large proportion of those GWAS traits were measured in mixed-sex samples, which suggests that traits with female-specific effects that are relevant for breast cancer risk could have been overlooked in the analysis (e.g. IGF-1 [44], testosterone [61]). Moreover, many known risk factors (from observational or previous MR studies) may not come up in the MR-EvE review for several reasons, (1) some traits are not heritable/instrumentable or not in OpenGWAS, (2) traits were measured in another (larger) cohort, or with additional covariates (e.g. BMI), (3) the filtering and FDR correction steps excluded traits with weaker evidence, which may have been reported by smaller studies (e.g. LDL-C).

Many traits identified using MR-EvE in this work have been published as individual MR studies over the recent years, despite MR-EvE data being available via EpiGraphDB since 2021 and earlier as a standalone resource (generated in 2017 by Hemani *et al* [14]) [82]. This highlights MR-EvE’s value in generating hypotheses and guiding analyses, while also stressing the redundancy in the growing number of MR studies, which often report only two-sample MR results without providing further evidence, or attempting to dissect the underlying biological mechanisms [83, 84].

In our study, we address the need for evidence triangulation in MR studies by incorporating literature-mined data to strengthen the evidence for identified relationships and to generate hypotheses for underlying mechanisms. To achieve this, we developed the LBD method (Figure 3) that helps to identify mechanistic pathways or mediators for causal associations by mining literature data in EpiGraphDB. The identified intermediate terms may then be tested using MR (given suitable GWAS data is available). In both presented case studies, a third of the identified intermediates provided testable hypotheses for MR validation. In this work, we limited GWAS data search to OpenGWAS due to practical constraints, but future research could extend this search to other sources (e.g. GWAS catalog [85]) and the newly released molecular datasets (e.g. proteomics data (Olink) [86] and lipids/metabolites data (Nightingale NMR) [87]).

LBD and MR can mutually enhance each other: LBD can complement MR causal inference by identifying potential intermediates, while MR validation of LBD findings helps address challenges in evaluating discoveries and benchmarking the performance of LBD methods [88, 18]. Without proper evaluation, it’s difficult to determine the usefulness or practicality of a discovery, which hinders the adoption of LBD in biomedical research [89]. Reviewing LBD discoveries with MR (if the terms are instrumentable and data is available) offers a relatively quick strategy for validation of the feasibility of the identified relationships. Certainly, MR is not a definitive answer for assessing traits’ causal associations or validating a potential mediation mechanism. However, applying it to LBD discoveries may help to prioritise the most plausible mechanisms, which could be taken further into more costly and time-consuming validation.

Despite the potential gains of incorporating LBD into the biomedical/ epidemiological research process, the limitations and pitfalls of working with literature-mined data should be noted. Even though literature space extraction from EpiGraphDB is a straightforward process, the underlying raw triples data is noisy and may be difficult to interpret. The extraction of semantic triples from text, mapping terms to UMLS and later to GWAS phenotypes, is challenging and could be imprecise, and therefore this data must be treated with caution. Additionally, it is important to note that the chains of intermediate terms (presented as Sankey diagrams) should not be interpreted as true biological pathways. They simply represent relationships between biological entities mentioned in publications, and therefore direct interactions cannot be assumed. Moreover, the extracted relationships may also be affected by publication bias (i.e. due to abstracts often emphasising the established knowledge). Therefore, any insights drawn from literature-mined data must be traced back to the original publications to verify the validity of extracted relationships before being used as evidence.

## 5. Conclusion

In this article, we presented a novel and efficient approach to undertake a systematic investigation of breast cancer risk factors by mining EpiGraphDB, representing the largest MR study of breast cancer so far. We also introduced a novel LBD method for identifying potential intermediates or mechanisms between traits of interest. Our data-driven approach highlighted that MR and LBD can be used as complementary methods, providing supporting evidence to each other, as well as guiding the generation of hypotheses and bringing evidence from observational and genetic sources together. However, further validation through sensitivity tests, MVMR, and mediation analyses (where appropriate) is essential, as these are important steps in validation of causal relationships. The risk factors and mediators identified in this work can be followed up in future studies in more detail, seeking evidence from other analyses or sources, such as observational studies or lab-based validation. Lastly, the overall approach can be used to study the aetiology of other diseases.

## 6. CRediT authorship contribution statement

**Marina Vabistsevits** - Conceptualization, Formal analysis, Data curation, Investigation, Methodology, Validation, Visualization, Writing – original draft, Writing – review and editing; **Yi Liu** - Supervision, Writing – review and editing, Methodology, Software; **Tim Robinson** - Supervision, Writing – review and editing; **Ben Elsworth** - Conceptualization, Supervision, Methodology, Software; **Tom R Gaunt** - Conceptualization, Supervision, Methodology, Software, Writing – review and editing, Methodology, Project administration, Funding acquisition

## 7. Declaration of Competing Interest

T.R.G receives funding from Biogen and GSK for unrelated research.

## Data Availability

All data produced in the present work are contained in the manuscript or publicly available.

## Acknowledgments

M.V. is supported by the University of Bristol Alumni Fund (Professor Sir Eric Thomas Scholarship). T.R. is supported by NIHR Development and Skills Enhancement Award (NIHR 302363) and has received grants to attend educational workshops from Daiichi-Sankyo and Amgen. M.V., T.R., Y.L., T.R.G, work in the Medical Research Council Integrative Epidemiology Unit at the University of Bristol supported by the Medical Research Council (MC UU 00032/03). This work was also supported by a Cancer Research UK programme grant (the Integrative Cancer Epidemiology Programme) (CC18281/A29019). This study was also supported by the NIHR Biomedical Research Centre at University Hospitals Bristol NHS Foundation Trust and the University of Bristol. The views expressed in this publication are those of the author(s) and not necessarily those of the NHS, the National Institute for Health Research or the Department of Health.

## Appendix A. Literature overlap method details

### Appendix A.1. Literature-mined relationships in EpiGraphDB - extended version

EpiGraphDB contains literature-mined relationships between pairs of terms (also known as ‘literature triples’) that were derived from the published literature. Figure A.5 summarises the tools/databases involved in making literature-mined data available in EpiGraphDB. Literature-mined relationships in EpiGraphDB were originally extracted by the MELODI-Presto tool [38]. The underlying literature data comes from SemMedDB (Semantic MEDLINE Database) [16], a well-established repository of literature-mined semantic triples, i.e. ‘*subject term 1 – predicate – object term 2*’, mined from titles and abstracts of nearly 30 million biomedical articles in PubMed, using SemRep [39, 40]. SemRep is a natural language processing (NLP) system that parses biomedical text using linguistic principles and UMLS (Unified Medical Language System) domain knowledge [90] to extract semantic triples.

**Figure A.5:**
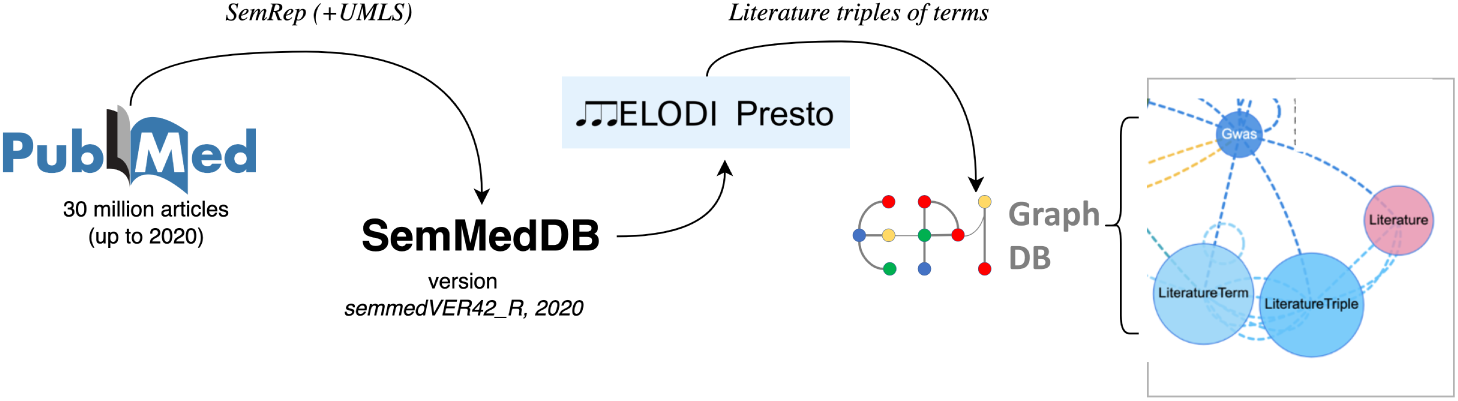
A workflow diagram showing the origins of literature data in EpiGraphDB and the tools/databases involved in making this data available. EpiGraphDB literature term/triples nodes (on the right) is a subset of the full EpiGraphDB schema: https://www.epigraphdb.org/about

### Appendix A.2. Literature space overlap method - extended version

Developing a method for connecting the literature spaces of exposure and outcome traits to explore potential links between them was one of the key aims of this work. The underlying principle of the literature space overlap approach is linking semantic triples of established biomedical knowledge into sequential ‘chains’, in order to uncover new connections. This principle of triple linking has previously been showcased in MELODI-Presto [38], and is an extension of the ABC paradigm [41, 43] with the expansion of a chain of Bs (*B*_1_ to *B_i_*): *AB^i^ C* (see 2.6.2). The details of the method are described below.

#### Triple cleaning

First, the extracted literature space for a trait is cleaned using several strategies – terms are standardised to minimise redundancy (i.e. merging long/short names, name acronyms, synonyms, gene/protein names); terms with different entity types (e.g. *aapp* - amino acid/protein, *gngm* - gene/genome) are filtered to include only the most common type; triples with the same *term1* and *term2* are excluded; among bidirectionally related terms the ones with more more common direction (indicated by triple score) is retained. Additionally, terms that represented drugs/medications or other diseases (except anthropometrics-related terms) as well as negative predicates (e.g. ‘*neg coexists with*’) are excluded. Finally, a ‘frequency score’ is generated for each pair of terms, i.e. the sum of triple scores across all possible predicates linking the terms, which is later used for triple chaining visualisation (Sankey diagrams).

#### Term anchoring

To implement *closed discovery* from the ABC paradigm [43] (see 2.6.2), we perform ‘term anchoring’, which involves restricting the outer term in a chain of triples to be the trait itself. For example, for an exposure trait (e.g. HDL-C), we are interested in downstream entities, so *term1* in the first triple of a chain make from its literature space would be *’HDL’* (Figure 3b: **“T”** for trait). Inversely, in the breast cancer literature space, we are interested in entities upstream of breast cancer, so we would restrict *term2* of the terminal triple in the chain to be *’Breast cancer’* (Figure 3b: **“BC”** for breast cancer).

For non-molecular traits, it is more difficult to perform term anchoring, because specific terms representing such traits are often not available. Therefore, for those we rely on *open discovery* [43] (see 2.6.2): the spaces are connected by matching unlinked triples from the exposure trait’s literature space to any breast cancer triples, and then extending chaining of the matched triples upstream into the trait’s space (Figure 3c).

#### Triple chaining

Literature spaces contain a variety of triples (some examples in Table A.5), and only a small proportion of them contain the trait itself as a term. Using only the triples that include the trait would make the analysis quite limited. Therefore, ‘triple chaining’ is implemented as a part of the approach to extend the discovery space. Within a trait’s literature space, we generate a basic set of triple chains, with the outer triple anchored to the trait term (see above), e.g. “**T**-A-B” and “C-D-E-F-**BC**”, where letters A-E are terms forming chains (see Figure 3b). Then, the sets of triples from two literature spaces are “overlapped”, which involves *term2* from the exposure trait’s space triples to match with *term1* in triples from the breast cancer literature space. This can be done at any possible point, i.e. not necessarily as the longest multiple-step chain (e.g. “T-A-B-C-D-E-F-BC”), but can be of any variation (e.g. “T-A-B-BC”, “T-D-E-F-BC”).

**Table A.5:**
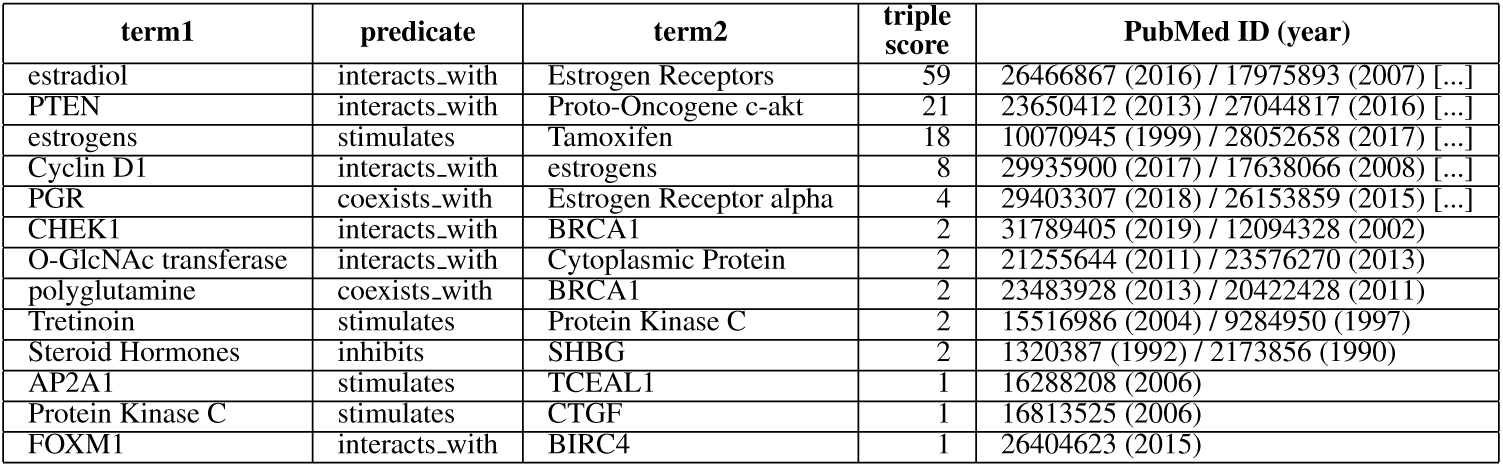
A subset of triples from the breast cancer literature space. (*term1 - predicate - term2*) (randomly selected for illustrative purposes). The triple score indicates the number of publications the triple appears in/was extracted from. The ‘PubMed ID (year)’ column contains (a subset of) PubMed IDs of those publications. Full data is in Supplementary Table 14.

#### Literature spaces

The literature overlap method performs best on literature spaces with *>* 50 triples, as this is required for effective triple chaining. Small literature spaces are also less likely to contain the trait as a term, which is required for term anchoring. Therefore, literature space sizes were also considered when choosing case study traits, to ensure an effective application of the literature overlap method. Literature spaces made of *<* 50 triples indicate that fewer publications were found to be linked to them. This may be due to the trait being less studied (and published about), or issues with the trait name (i.e. non-standard phasing of a concept), or issues with the source data in SemMedDB [16] or trait mapping in MELODI-Presto [38] (Figure A.5). Literature spaces of this size are generally unusable in the literature space overlap method.

### Appendix A.3. Literature spaces - extended details

#### Breast cancer

Breast cancer literature space was formed from data linked to two traits: ‘*Breast cancer*’ (OpenGWAS ID: *ieu-a-1126*) and ‘*Malignant neoplasm of the breast*’ (*finn-a-C3 BREAST*). 18,848 unique triples were identified, based on 4989 unique terms derived from 23,809 publications. The clean literature space is available in Supplementary Table 14. The breast cancer anchor term is ‘*Breast cancer*’, which is combined from the terms ‘*breast diseases*’ and ‘*malignant disease*’.

#### Case studies

The selected case studies include *HDL* (*’HDL-cholesterol’* - *ukb-d-30760 irnt*, 7808 unique triples) and *childhood body size* (*’childhood obesity’*-*ieu-a-1096*, 1255 unique triples) (Supplementary Tables 15-16). For childhood body size, we chose to use *’childhood obesity’* literature space over *’Comparative body size at age 10’* (*ukb-b-4650*, 211 triples) due to the larger space size and trait name specificity.

### Appendix B. Sankey diagrams for case studies

**Figure B.6:**
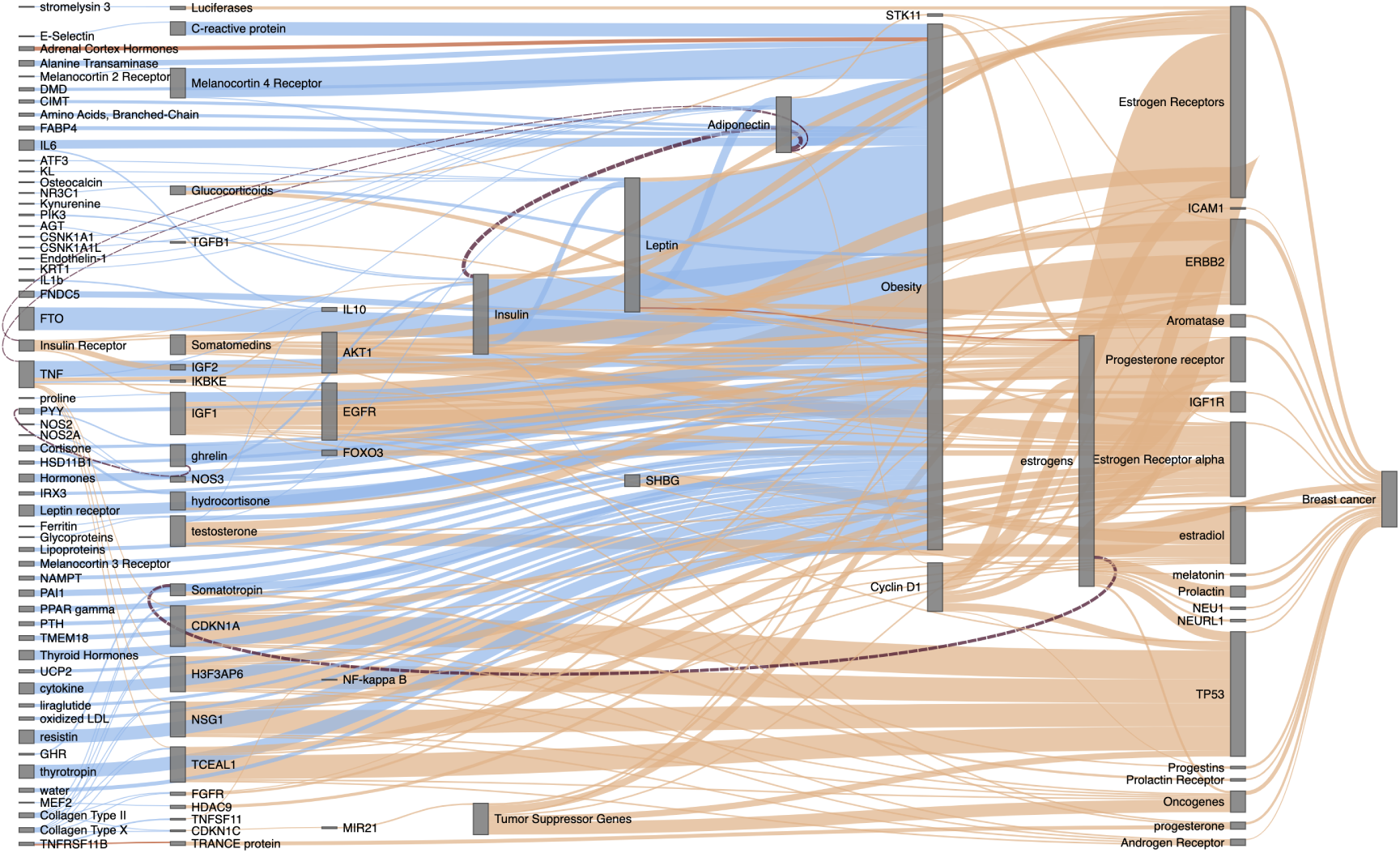
Sankey diagram of childhood obesity and breast cancer literature spaces overlap, showing the overlap of literature triples within and between the spaces. The blue and orange relationships (triples) come from the childhood obesity and breast cancer literature spaces, respectively. The line width of each term relationship indicates how common it is in the literature (frequency score). Here, the open discovery approach was used, i.e. only one side of the overlap is anchored (*’Breast cancer’)*; View interactively at https://mvab.shinyapps.io/literature_overlap_sankey/

**Figure B.7:**
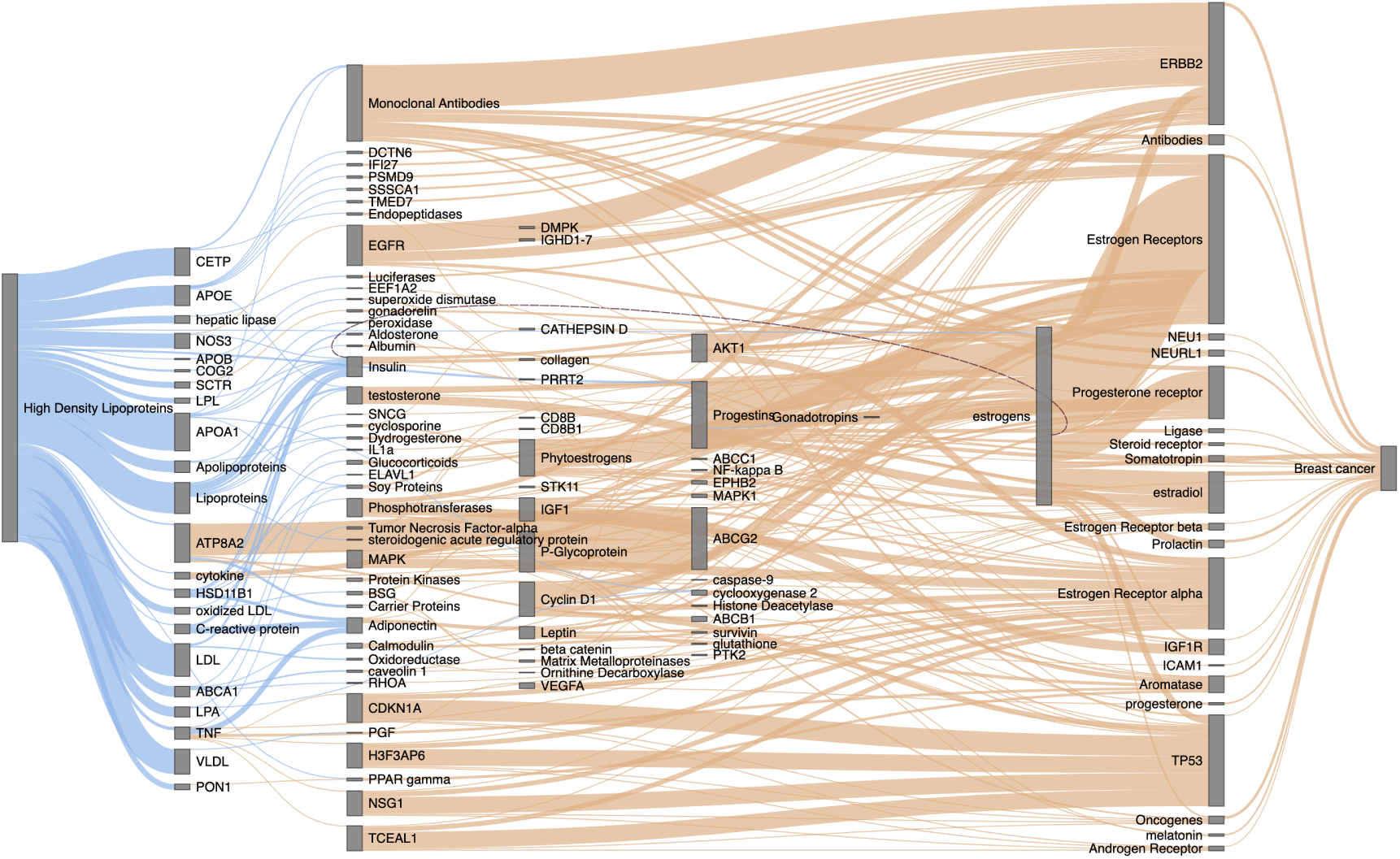
Sankey diagram of HDL and breast cancer literature spaces overlap, showing how triples within and between HDL and breast cancer spaces interconnect. The blue and orange relationships (triples) come from the HDL and breast cancer literature spaces, respectively. The line width of each term relationship indicates how common it is in the literature (frequency score). Here, the closed discovery approach was used; the anchor terms are ‘*High Density Lipoproteins*’ and ‘*Breast cancer*’. View interactively at https://mvab.shinyapps.io/literature_overlap_sankey/

### Appendix C. Extended discussion of MR results for dietary and physical traits

The final set of 129 validated breast cancer risk factors identified from MR-EvE included several dietary traits. Genetically-proxied dietary intake exposures require careful consideration due to the distinct sources of bias and interpretation challenges they present. Dietary intake GWAS in UK Biobank can produce a considerable number of instruments due to their sample size, and the risk of horizontal pleiotropy among these SNPs is high [91]. Trait heterogeneity is also important to consider [92], i.e. the fact that genetic instruments associated with the dietary trait may represent different ‘dimensions’ of the same trait, e.g. intake of a certain food item and metabolism of its core component (e.g. coffee/caffeine [93]). The primary risk in over-interpreting dietary intake MR lies in the possibility that the extracted instruments may be non-informative or associated with dietary patterns through socio-demographic and behavioural factors, potentially confounding the MR results [91]. Additionally, dietary trait instruments often include adiposity-related signals, meaning these variants could be affecting dietary patterns via their association with adiposity, leading to this association being captured in the MR results.

In this study, dietary traits with the most consistent evidence of effect were *‘dried fruit intake’* (showing a protective effect) and *‘processed/sugary cereal intake’* (associated with an increased risk). Despite those MR results showing good instrument strength and no strong indication for pleiotropy but some heterogeneity, we cannot claim that the captured effect is specific to those food items. The traits could be proxying two different environmental exposures, e.g. related to a broader class of foods that share similar characteristics, or overall dietary pattern, i.e. ‘healthy’ and ‘unhealthy’ food choices. The “gene-environment equivalence” assumption in MR states that the downstream effect of exposure modification is the same, regardless of it being genetically or non-genetically triggered. It could be hypothesised that the personal preference/choice to consume dried fruit vs sugary/processed cereal could be reflective of the other dietary patterns in those individuals’ respective diets. Overall, the effect direction matches with the observational evidence of diet impact on breast cancer risk [94]. The ‘healthy’ dietary pattern includes a varied diet with high fruit and vegetable intake, which increases the overall fibre and micronutrient content in the diet (more likely to occur in those consuming dried fruit), while the ‘unhealthy’ pattern involves a larger amount of processed and less nutrient-rich food items (more likely to occur in those choosing to eat sweetened breakfast cereals). The genetic correlation analysis in the study by Pirastu *et al* [95] has shown a positive correlation of ‘dried fruit intake’ GWAS with other traits relating to a healthier diet (e.g. intake of fresh fruit, cooked veg, fish, salad, being vegetarian) and a negative correlation with dietary patterns considered less healthy (e.g. alcohol intake, processed meat, beef, spread on bread, salt, instant coffee), supporting the idea that *‘dried fruit intake*’ instruments may be proxying an overall healthier diet. The protective effect of ‘*dried fruit intake*’ on breast cancer discovered from MR-EvE has been recently published in a separate MR study [96], in which the authors also reviewed the most likely confounders of this relationship (e.g. vitamin C, BMI, years of education) using MVMR, and found that the identified inverse relationship is not affected when adjusted for any of them.

*Skimmed milk intake* was another dietary trait identified from MR-EvE with evidence of a protective effect on breast cancer, but this result has to be carefully considered as it is misleading. Previous MR studies of milk intake effect [68, 69] selected instruments (nSNP=1) specifically within the lactase gene locus (*LCT*), which encodes an enzyme that ensures tolerance of milk products. The variation in this locus affects the amount of milk consumed and, therefore, is often used as a proxy for this exposure. Both MR studies found a mild risk-increasing effect from milk intake. In MR-EvE data, the instruments for *‘skimmed milk intake’* trait were extracted genome-wide (nSNP=3), none of them being in *LCT*. MR sensitivity tests identified likely horizontal pleiotropy and heterogeneity. This serves as a valuable negative example of dietary traits evaluation and highlights the importance of conducting more detailed investigations of these traits in individual studies.

Physical activity is one of the modifiable lifestyle factors that have been shown to have a protective effect on breast cancer risk, particularly vigorous activity [97]. MR studies using accelerometer-measured overall physical activity produced results to support this [70, 50]. Among the physical activity traits available in MR-EvE (UK Biobank traits based on a survey question *“Types of physical activity in last 4 weeks”*), several showed similar protective effects: *‘Heavy DIY’, ‘strenuous sports’* and *‘exercise to keep fit’* (the last identified in a case study), which may be proxying moderate-to-vigorous physical activity. Similarly, there was a risk-increasing effect from *‘no physical activity’* trait, which is in agreement with ‘sedentary time’ found to increase the risk in [50] and the observationally-known detrimental effects of physical inactivity [98]. With survey-derived physical activity phenotypes (such as *‘Heavy DIY’*) appearing unreliable, a potential follow-up to these findings would be to compare the genetic instruments identified for physical activity from survey phenotypes to those derived from accelerometer-measured data.

## Notes

### Author Declarations

The study used only openly available human data.

### Summary of Updates

The revised version of this manuscript is more concise and focuses on just two case studies. The updated discussion addresses the current issues with MR studies publishing, which is directly relevant to this MR Everything-vs-Everything study.

